# Theoretical Basis for an Edge-based, mHealth App to Guide Self-Management of Recurrent Medical Conditions

**DOI:** 10.1101/2020.04.28.20082339

**Authors:** Alexander M. Kaizer, Susan L. Moore, Farnoush Banaei-Kashani, Sheana Bull, Michael A. Rosenberg

## Abstract

**Background:** N-of-1 trials have been proposed as an approach to identify the optimal individual treatment for patients with a number of recurrent medical conditions, including chronic pain and mental health. When inserted into mHealth applications, this approach holds great promise to provide an automated, efficient method to individualize patient care; however, prior to implementation, an understanding of the properties of the recurrent condition needed to draw conclusions with sufficient power is needed.

**Methods:** We applied simulation studies and power calculations to determine statistical properties of the N-of-1 approach employed by an mHealth application for self-management of chronic recurrent medical conditions called the iMTracker.

**Results:** In 1000 simulated patients with a single recurrent medical condition and 5 possible associated conditions, we found that ~90 days of data collection was sufficient to identify associated risk factors with odds ratio (OR > 5.0) at power ≥ 80%, with an absolute event rate of 50% being optimal. Power calculations based on Fisher’s Exact test showed that 90 days was also sufficient to detect a decrease of 20% in the rate of the primary outcome after an intervention, but that shorter data periods could be used to identify stronger effect sizes, down to 15 days with a 90% reduction in rate. Repeat analysis with Bayesian models did not significantly change power calculations, but did allow for a flexible approach that we leveraged to create a web-based tool to allow users to perform power calculations prior to using the iMTracker for self-management.

**Conclusions:** We found that the N-of-1 approach employed in the iMTracker app for self-management of recurrent medical conditions is statistically feasible, given the right conditions. More work is needed to examine the impact of autocorrelation, seasonality, and trends in data, on statistical validity and power calculations.

## Background

Management of chronic recurrent medical conditions, such as low back pain or migraine headaches, presents a major challenge for the current time-constrained healthcare system. Although evidence-based guidelines have been developed for management of these conditions across entire populations^1–5^, application to individual patients is often much more challenging. For example, sleep changes have been described in ~50% of patients with migraine headaches, although 75% of patients also chose to sleep due to the migraine headache itself^6^. In general, the approach to management of most patients with these conditions involves the patient providing a hand-written journal or log of the daily presence of recurrences at the time of a 20–30 minute clinic visit; a terse, qualitative review of the pattern by the provider searching for obvious patterns; a brief discussion of possible triggers, suppressors, or response to treatment using subjective language (e.g., ‘Does caffeine help/hurt?’); and a nonsystematic plan to make a series of uncoordinated changes in lifestyle or medications, followed by scheduling of a follow-up visit at some arbitrary timepoint to assess the impact. The motivated patient may continue to log the recurrences until follow-up, at which time the exercise is repeated, generally with subjective assessments of results.

The motivation for N-of-1 clinical trials is to develop a methodology for systematic comparison of treatments or exposures on an outcome at the individual patient level. The N-of-1 approach has been applied to study various interventions for pain^7–11^, depression^12–14^, anxiety^15, 16^, and migraine headaches^17–19^, as well as countless applications within the clinical realm that are unreported in the literature due to the challenges with extrapolation to the greater body of evidence^20^. Noteworthy, and perhaps characteristic, of many N-of-1 trials is that they are generally are limited to only a few patients—from one to tens of patients, reflective of the practical demands of manual data collection and analysis on a per-person basis. To date, no large clinical trial has been performed using the N-of-1 approach that has demonstrated an improvement in clinical outcomes, which highlights both the challenges and opportunities to apply technology innovations toward patient care.

The possibility that technology could improve individualized patient care, with or without the specific N-of-1 approach, has been raised by providers and innovators alike. Patients are motivated, and will often seek advice or guidance outside the realm of expert clinicians. As a result, the market for so-called self-management applications and nonstandard care approaches is flooded with untested and unvalidated smartphone apps and clinics that promise patients the opportunity to self-manage their conditions. Like many technology innovations, the companies providing these solutions often focus more on commercialization and marketing rather than scientific assessment of their applications^21–26^, leaving both providers and patients skeptical about incorporation into clinical care.

To meet this emerging need, our team of clinicians, statisticians, and investigators have developed an iOS application called the iMTracker, which is currently available for nonmedical use on the iTunes store^27^. Starting with a broad approach focused on the identification of lifestyle factors (e.g., caffeine) that could be associated with recurrence of a chronic condition (e.g., migraines), our approach includes a process to test any intervention (e.g., drink less caffeine) that the user would like to test on the recurrence pattern of the outcome. Through iteration between hypothesis generation (i.e., ‘is there an association between risk factor A and occurrence of my condition?’) and hypothesis testing (i.e., ‘does changing risk factor A improve the rate of occurrence of my condition?’), the user is able to self-manage his or her condition towards an overall goal of reducing recurrence.

However, before such an approach can be examined in large clinical trials, there are key questions about the statistical validity of this process to identify triggers and/or suppressors for a given patient’s recurrent condition. Specifically, how much data are needed—how often and for how long does the patient need to collect data—in order to draw inferences with sufficient statistical power to guide care decisions? In this study, we examined the impact of changes in the recurrence rate of the primary condition, the rate of possible triggers or suppressors, and the strength of association between these factors, on the statistical power of this approach to guide care decisions. We start with an assessment of the approach of collecting data on possible associations to identify possible triggers or suppressors, for future interventions, and then examined the amount of data to determine the effect of a chosen intervention on the recurrence rate of the outcome of interest. Throughout, we make the assumption that simpler statistical models that can be easily programmed into a mobile device (without need to transfer data to a separate server) will be preferred over more complex approaches that would require greater computing power or incorporation of data from other users.

## Methods

### N-of-1 Paradigm

The N-of-1 paradigm used in this investigation is shown in Figure 1. Essentially, the patient is prompted to enter the presence or absence of a given condition, as well as a selected number of possible triggers or suppressors (Factor X, Fig. 1), on each day for the duration of the data collection period. After a determined data collection period (N days), the association between the possible trigger or suppressor is calculated as the odds ratio of the association, which is reported back to the patient along with the strength of the association. Based on the strength of the association, the patient would be advised to either 1) avoid the factor (OR > 1, indicating a possible trigger), 2) seek out the factor (OR < 1, indicating a possible suppressor), 3) select alternative possible factors due to a sufficiently powered lack of effect (OR ~1, power > 0.8), or 4) continue to collect data due to lack of power to make a determination (Power < 0.8).

**Figure 1.**
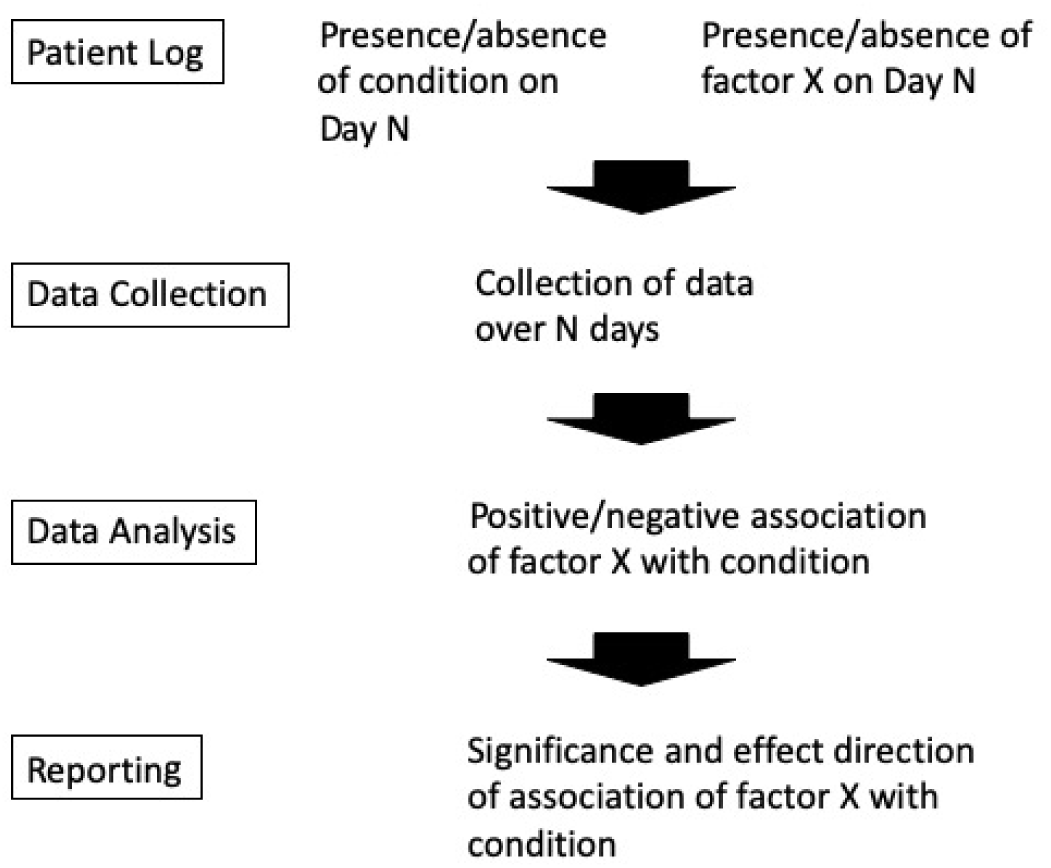
Outline of the process of self-management used by the iMTracker.

### Simulation Parameters

Based on the above model paradigm, we identified the following parameters to examine using simulation:

N = Number of days to collect data
Power = 1 – false negative probability (probability of a false null association)
P(Event) = Probability of having the recurrent event
P(Factor) = Probability of exposure to the factor
P(Factor|Event) = Probability of having the factor present on the day of a recurrent event
P(Factor| No event) = Probability of having the factor present on a day without the recurrent event
OR = Odds ratio between factor and recurrent event

### Simulation Model

The simulation approach employed was guided by the principle that patients will prefer to collect data for the minimum amount of time needed to draw an association with sufficient statistical power to identify a ‘positive’ (OR ≠ 1) association or ‘null’ (OR = 1) association. Simulation studies were therefore used to evaluate the power or type I error rate for alternative (i.e., ‘positive’) or null scenarios, respectively. The existing simulation studies are all completed in R (version…) with 1000 simulated people experiencing 5 independent triggers with different probabilities of the triggers.

We simulated a total of 180 days of data with no missed entries assumed (i.e., the participant always enters their data to the iMTracker app). Summaries are provided for statistical properties at days 7, 14, 28, 60, 90, 120, and 180. The current scenarios examined include assuming a 20%, 50%, and 80% event rate for the following probabilities of trigger exposure (Table 1). To examine the role that more permissive statistical criteria for possible associations could be identified by relaxing the Type I error rate (alpha, significance threshold), we also repeated simulations using an alpha cut-off (probability of falsely rejecting null hypothesis) of 0.05. We then examined the role of time of data collection using the log of the odds ratio, as early simulations found that it better represented the estimates. Negative values indicate OR<1 and positive values indicate OR>1. Estimates of OR=0 or OR=infinity were excluded to avoid calculation issues, and were denoted as “Conditional ORs”.

**Table 1.** Simulation Parameters.

| Scenario Type | P(Trigger\|Event) | P(Trigger\|No Event) | Odds Ratio |
| --- | --- | --- | --- |
| Null Scenarios (Scenarios with OR=1) | 0.20 | 0.20 | 1.0 |
|  | 0.35 | 0.35 | 1.0 |
|  | 0.50 | 0.50 | 1.0 |
|  | 0.65 | 0.65 | 1.0 |
|  | 0.80 | 0.80 | 1.0 |
| Alternative #1 (Scenarios with OR>1; Trigger is Harmful) | 0.25 | 0.20 | 1.3 |
|  | 0.40 |  | 2.7 |
|  | 0.55 |  | 4.9 |
|  | 0.70 |  | 9.3 |
|  | 0.90 |  | 36.0 |
| Alternative #2 (Scenarios with OR<1; Trigger is Protective) | 0.20 | 0.25 | 0.75 |
|  |  | 0.40 | 0.38 |
|  |  | 0.55 | 0.20 |
|  |  | 0.70 | 0.11 |
|  |  | 0.90 | 0.03 |

### Intervention Effect Analysis

Once a hypothesized association has been identified, the critical step is to then test whether the intervention provided a statistically significant change in the outcome of interest. Such an assessment requires a minimal amount of data to be collected before and after the intervention, which depends on the size of the effect of the intervention on frequency of the recurrent outcome. To examine the amount of data needed, we performed power calculations using Fisher’s Exact test of proportions (Stata, IC., version 15, StataCorp, Inc., College Station, TX, USA). We examined data collection periods from 10 to 100 days (pre-intervention), with the goal of identifying interventions resulting in a decrease in the frequency of the recurrent event at significance level (alpha) of ≤ 0.05 and power ≥ 0.8.

In appreciation that real-time use of the iMTracker would entail online analysis during data collection, we also compared use of a Bayesian model based on a Beta distribution for the distribution of the proportion of events/total days as such: Pr(y) ~ Beta(a, b), where a = # of events + 1 and b = total days − # of events + 1.

This Bayesian tool was then coded into a web-based analytical tool using dash v1.4.1 (https://dash.plotly.com), hosted on Heroku (https://heroku.com). The analysis was performed using Python, v 3.7.4, including scipy.stats.beta (v1.4.1), numpy v1.18.1, Flask v1.1.2, matplotlib v 3.1.3 (see Supplemental material for source code).

## Results

We first verified that the simulation was insensitive to null scenarios, OR = 1.0, and that the type 1 error rate was maintained across all null scenarios with all event rates, never exceeding 5% in any case and being lower with fewer days of data, at all event probabilities (Supplemental Fig. 1A-1C). We then modeled simulations for various event probabilities, based on different probabilities for no event vs. event (see Table 1 for details), and found that as the OR increases, the power increases for a given number of days. We found that for a very low OR, where the trigger is only marginally higher for the event vs. nonevent days (i.e., 25% vs. 20%), we never achieved power above ~7% at 180 Days. Further, we found that an event rate of 50% was optimal to maximize power, because it maximizes our information about event and non-event days and if the trigger is experienced (Figures 2A-C).

**Figure 2.**
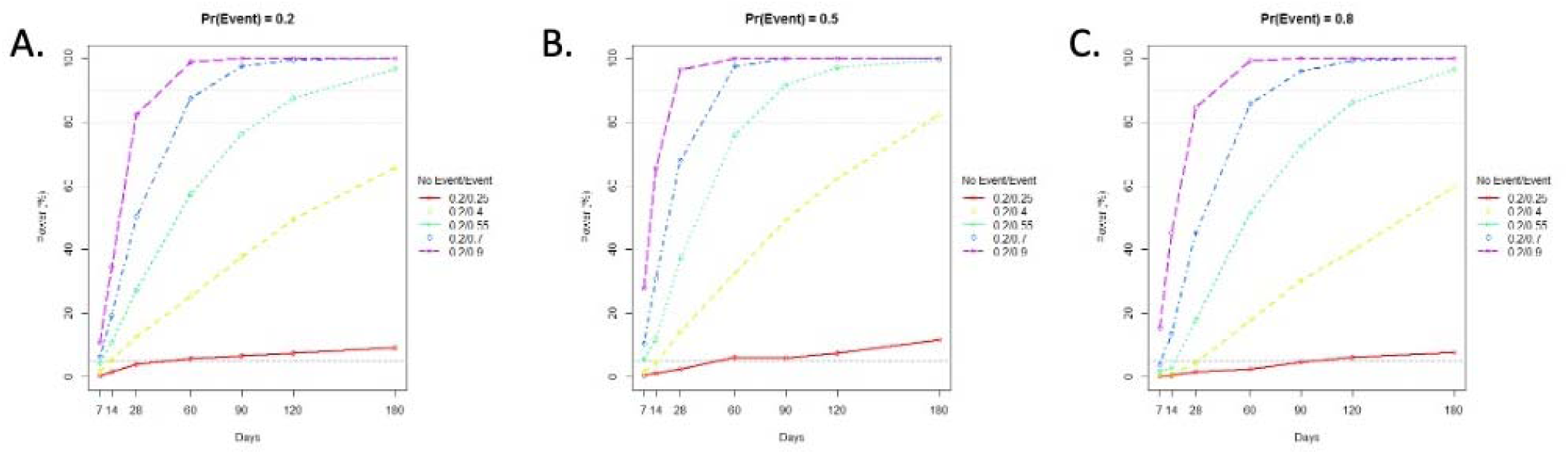
Simulation Results for Possible Triggers (i.e., OR>1) with P(event) = 0.2 (A), 0.5 (B), and 0.8 (C)

To validate that effects could be detected in both directions (both triggers and suppressor associations), we repeated this analysis by keeping P(trigger|event) at 0.2, and increasing the P(no trigger|event), which also showed that P(event) of 50% was optimal, and that OR < 0.4 could not be detected with 80% power at any time period of collection (Supplemental Fig. 2A-2C). Relaxation of the significance threshold of events to alpha = 0.2 had a minor effect on power (Supplemental Fig. 3A-3C), although less than increasing duration of data collection. This characteristic was validated in additional null simulations, which found that in general, the mean log(OR) converges quickly to a value of 0 (i.e., the null). Noteworthy, we found that between 76.6% to 91.3% of estimated OR are 0 or infinity at day 7, between 41.5% to 66.6% at day 14, between 9.9% to 35.5% at day 28, and still occurring intermittently by chance at later days (Supplemental Fig. 4A-4C). By excluding OR estimates of 0 or infinity, we found that the remaining estimates converged to the true mean log(OR) value, although these estimates generally underestimated the true log(OR) at smaller numbers of days. However, they generally converge to the estimated OR near day 28 and more definitively at day 60. By day 60, the mean was fairly consistent, with fewer numbers of days having underestimates of the relationship (Supplemental Figs. 5 and 6). Broadly, these results demonstrated that the most stable estimates of effect required at least 60 days of data collection, but that 90 days of data collection provided the optimal duration to identify most associations with OR > 5.0, and that even 180 days of data collection could not identify lower ORs, with the exception of an event rate (P(event)) of 50%.

We then examined the impact of the amount of data collection (days) before and after the intervention in order to detect a significant change in frequency of the recurrent event using Fisher’s exact test. Assuming equal periods of time before and after the intervention, we found that regardless of the event frequency, no significant change could be detected with data collection under 15 days. As shown in Figure 3, for increasing numbers of days with data collected, the proportion of days with events and the relative reduction in frequency of events decreases to detect a significant change as a result of an intervention. For example, with only 15 days of data collected, an event would need to occur on minimum of 55% of days (~8 events) and have a reduction of at least 90% (~1 events) after the intervention, or an event could occur on 90% of days (~13 events) with a decrease of 60% (~8 events). However, with longer data collection, less frequent events and lower proportionate reduction can be detected.

**Figure 3.**
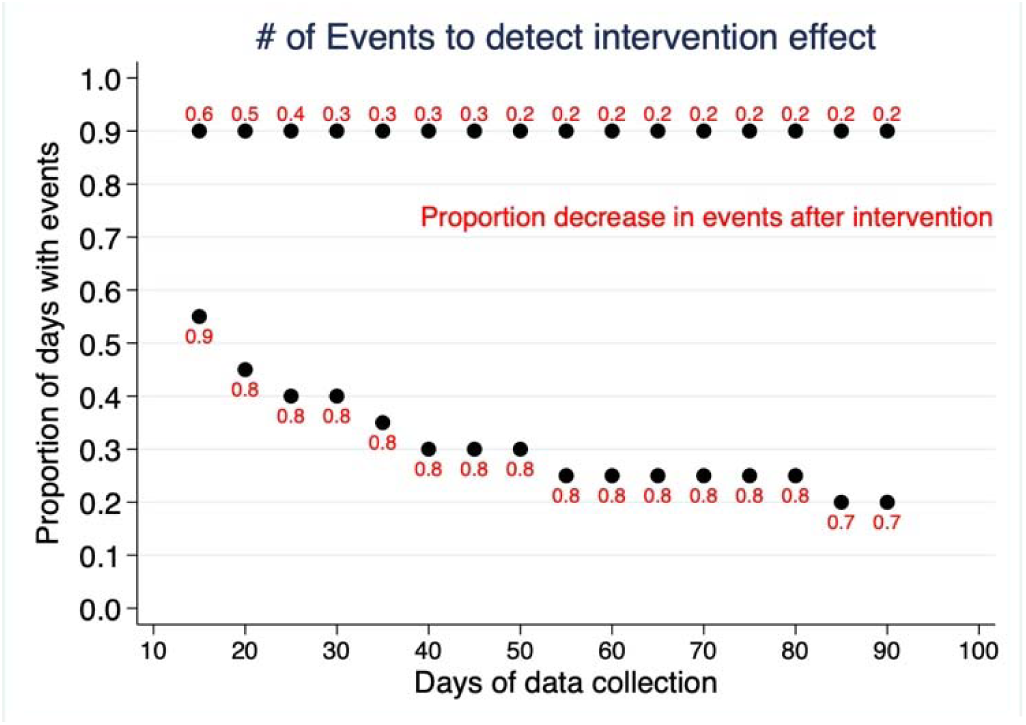
Intervention Effect. Red labels indicate the proportionate decrease in events after the intervention, compared with prior to intervention.

Finally, to examine the use of Bayesian statistics to compare interventions, we compared several effect rates over varying data collection periods and found that for event rates < 20%, only very high effect sizes could be detected with probability of 95% (Figure 4, Supplemental Figs. 6 and 7). To allow potential app users to examine their own data requirements, we developed a web-based app that is available at https://imtracker-power-calc.herokuapp.com (Supplemental Fig. 8).

**Figure 4.**
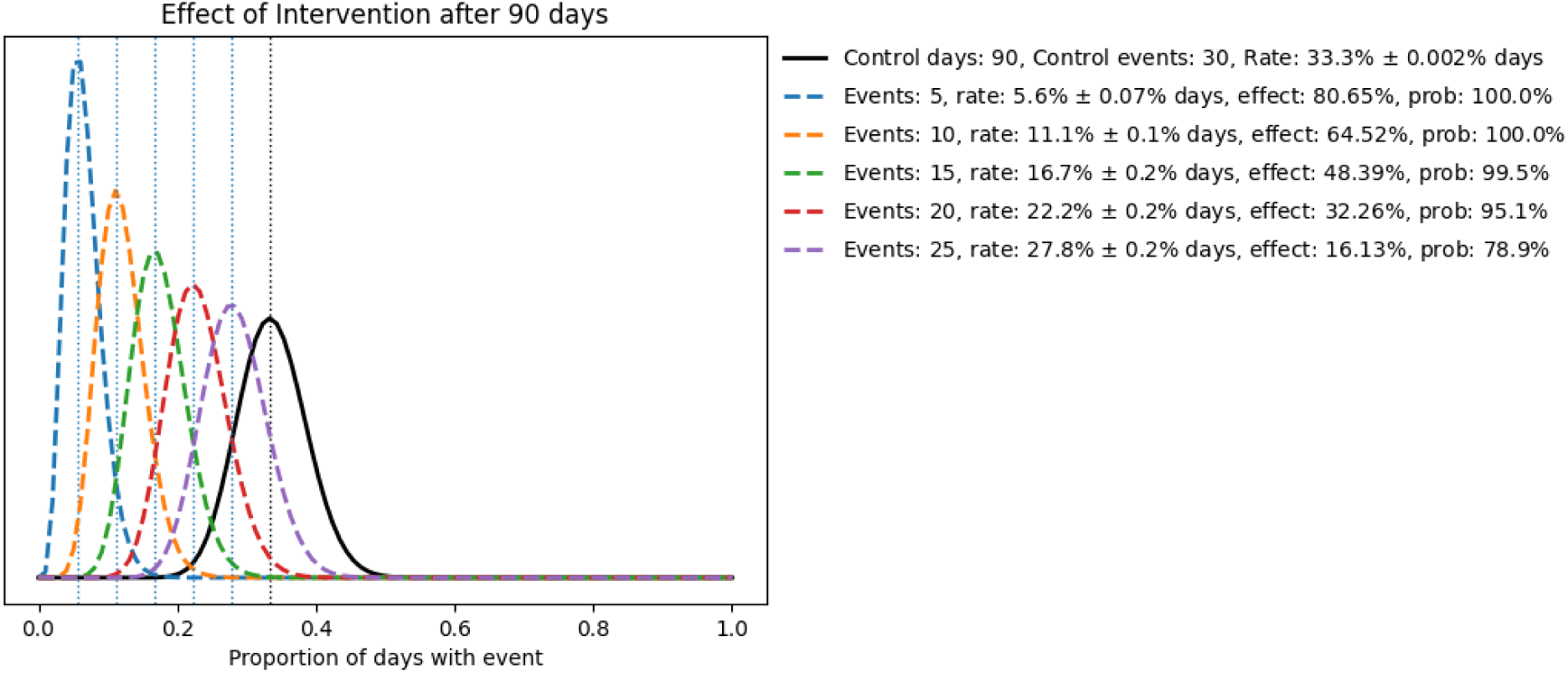
Bayesian effect calculations. Based on 90 days of data collection during the ‘control’ period, with rate of 33% (30 events). Comparisons include interventions with effect size ranging from 16.1% to 80.7%. Probability based on overlap of 95% credible intervals for the probability distribution of each group.

## Discussion

In this simulation study of the N-of-1 approach used by the iMTracker mHealth application to guide self-management of recurrent medical conditions, we identified several important characteristics about the duration of data collection needed, the association of possible risk factors (triggers or suppressors) for a given outcome, and the potential of an intervention to reduce the rate of recurrence of the primary outcome. First, we found that in terms of identifying associations between triggers or suppressors and a recurrent outcome, an outcome that occurred with probability of 50% was optimal for identification of associated factors, and that most associations of sufficient effect size to be identified with 80% power were seen by 90 days, indicating that there is little evidence for extending the data collection period beyond 90 days, at least for the purposes of identification of possible associations for intervention. In terms of testing interventions, we found that there was an inverse relationship between the intervention effect and the number of days needed to collect and test data to compare, with 90 days again being optimal in terms of providing sufficient power to detect an effect as low as 20%, albeit with an increased number of events needed during the data collection period.

These results have implications both for the specific usability of the iMTracker as a self-management app, as well as the world of N-of-1 studies broadly. For one, our study provides a general timeline for data collection of 90 days, during which the app can both identify possible associated risk factors for modification, as well as establish a baseline event rate for evaluation of future interventions. We note that event rates down to 20% can be reasonably quantified during this time period, acknowledging that any intervention would need to have relatively a strong association/effect to detect an improvement if targeted as an intervention. We found that fairly large odds ratios (~5) are needed to identify an association with sufficient statistical power, and little is gained by lowering the significance threshold needed to identify associations compared with increasing duration of data collection.

To guide this process, we also released an online, web-based power calculator that patients and providers can apply prior to use of the iMTracker to determine if the frequency of the event, and planned data collection period, will be sufficient to identify a change from the intervention with sufficient probability. Such information is valuable, as it can directly inform patients that an event that is uncommon, or for which proposed interventions would be inferred to have mild effects, is unlikely to meet criteria for use of the iMTracker in self-management. In contrast, for patients with frequent events, patients and providers would have the opportunity to develop a specific data collection plan, with details about exactly how long the app will need to be used.

Compared with standard clinical approaches, in which population-level data can be analyzed for the average treatment effect, N-of-1 medicine requires a highly automated process to scale, an area uniquely suited for advances in mHealth technologies. There is early evidence that an individualized approach to self-management of CRMCs using mHealth applications has potential to improve clinical outcomes for CRMCs. Kravitz et al., examined an mHealth-supported N-of-1 trial vs usual care in 215 patients with chronic musculoskeletal pain over 2 years, and found that 88% of those in the N-of-1 group affirmed that the mHealth app could potentially help manage their pain^28^, although the trial outcome found no significant difference between groups. Pumbo et al., examined 50 studies of mobile apps for pain management, and found data to be as accurate and feasible as pen-and-paper methods for symptom tracking, but noted that data integration presented a critical barrier to development and application of these approaches^25^.

We developed the iMTracker app out of frustration over the lack of an established quantitative methodology for assessment of patterns of recurrence of recurrent medical conditions, and response to treatment. In exploring N-of-1 approaches it became evident that to study the broader clinical impact of such an approach using randomized controlled trials would require a level of scale that was not feasible with manual application. Through use of an automated, algorithmic approach to management of recurrent medical conditions, we have identified both great potential and challenges. One particular challenge was concerns over data security and loss of privacy^31, 32^ that come with storing and analyzing data on a server, which can be compromised. In contrast, we designed the iMTracker based on the goal of using edge computing^29, 30^ strategies that run on the mobile device itself, in order to allow complete usage of the iMTracker without need for transfer or storage of data on a server. Although such features are desirable from a user perspective, there are challenges in attempting to move the entire data analysis platform to the edge, rather than maintaining data and data analysis on a server platform. For one, more advanced data analysis techniques, such as those integrating probabilistic programming like Markov chain Monte Carlo sampling for more advanced Bayesian analysis, are highly limited due to the memory requirements. Our Bayesian model employs the conjugate priors of the Beta distribution to allow quick analysis without complex computing. However, there are multiple simplifying assumptions in these analyses that could hold prominent effects in real world applications. Most specifically, these methods assume independence of events, and do not account for the dynamic, temporal nature of data collection, which includes both seasonal/trend effects, as well as intra-individual correlation (autocorrelation). These attributes can be modeled using more advanced approaches, such as using hidden Markov models, although at the expense of increased computational demands. Additional approaches, using federated learning to allow one user’s data to inform another are also possible, although these require more advanced networking technologies, an area of active research by our team.

In conclusion, we found that in general, event rates of 20% with data collection period of 90 days provides adequate power to identify possible associated risk factors, and test the impact of interventions. We provide an online power calculator for users to conduct their own power calculations, which through use of conjugate priors allow small computational demands as can eventually be included in the app logic itself. Future work on edge-based computing and federated learning is likely needed to improve the ability of these models to account for time-series features, such as autocorrelation and seasonality and trends.

## Data Availability

Data will be made available upon request and approval by co-authors

## Acknowledgements

This research was supported by grants from the NIH/NHBLI (MR: K23HL127296).

## Notes

### Competing Interest Statement

The authors have declared no competing interest.

## References

1. Practice guideline update summary: Pharmacologic treatment for pediatric migraine prevention: Report of the Guideline Development, Dissemination, and Implementation Subcommittee of the American Academy of Neurology and the American Headache Society. Neurology. 2020;94:50.

2. Leung A, Shirvalkar P, Chen R, Kuluva J, Vaninetti M, Bermudes R, Poree L, Wassermann E, Kopell B and Levy R. Transcranial Magnetic Stimulation for Pain, Headache, and Comorbid Depression: INS-NANS Expert Consensus Panel Review and Recommendation. Neuromodulation: journal of the International Neuromodulation Society. 2020.

3. Probyn K, Bowers H, Mistry D, Caldwell F, Underwood M, Patel S, Sandhu HK, Matharu M and Pincus T. Non-pharmacological self-management for people living with migraine or tension-type headache: a systematic review including analysis of intervention components. BMJ open. 2017;7:e016670.

4. Ng JY and Mohiuddin U. Quality of complementary and alternative medicine recommendations in low back pain guidelines: a systematic review. European spine journal: official publication of the European Spine Society, the European Spinal Deformity Society, and the European Section of the Cervical Spine Research Society. 2020.

5. Meyer T and Wulff K. Issues of comorbidity in clinical guidelines and systematic reviews from a rehabilitation perspective. European journal of physical and rehabilitation medicine. 2019;55:364–371.

6. Kelman L and Rains JC. Headache and sleep: examination of sleep patterns and complaints in a large clinical sample of migraineurs. Headache. 2005;45:904–10.

7. Notcutt W, Price M, Miller R, Newport S, Phillips C, Simmons S and Sansom C. Initial experiences with medicinal extracts of cannabis for chronic pain: results from 34 ‘N of 1’ studies. Anaesthesia. 2004;59:440–452.

8. Germini F, Coerezza A, Andreinetti L, Nobili A, Rossi PD, Mari D, Guyatt G and Marcucci M. N-of-1 Randomized Trials of Ultra-Micronized Palmitoylethanolamide in Older Patients with Chronic Pain. Drugs Aging. 2017;34:941–952.

9. Joy TR, Monjed A, Zou GY, Hegele RA, McDonald CG and Mahon JL. N-of-1 (single-patient) trials for statin-related myalgia. Annals of internal medicine. 2014;160:301–310.

10. Green AL, Shad A, Watson R, Nandi D, Yianni J and Aziz TZ. N-of-1 Trials for Assessing the Efficacy of Deep Brain Stimulation in Neuropathic Pain. Neuromodulation. 2004;7:76–81.

11. Yelland MJ, Poulos CJ, Pillans PI, Bashford GM, Nikles CJ, Sturtevant JM, Vine N, Del Mar CB, Schluter PJ, Tan M, Chan J, Mackenzie F and Brown R. N-of-1 randomized trials to assess the efficacy of gabapentin for chronic neuropathic pain. Pain Med. 2009;10:754–761.

12. Kronish IM, Hampsey M, Falzon L, Konrad B and Davidson KW. Personalized (N-of-1) Trials for Depression: A Systematic Review. J Clin Psychopharmacol. 2018;38:218–225.

13. Jansen IH, Olde Rikkert MG, Hulsbos HA and Hoefnagels WH. Toward individualized evidence-based medicine: five “N of 1” trials of methylphenidate in geriatric patients. Journal of the American Geriatrics Society. 2001;49:474–476.

14. Cook DJ, Guyatt GH, Davis C, Willan A and McIlroy W. A diagnostic and therapeutic N-of-1 randomized trial. Can J Psychiatry. 1993;38:251–254.

15. Malboeuf-Hurtubise C, Lacourse E, Herba C, Taylor G and Amor LB. Mindfulness-based Intervention in Elementary School Students With Anxiety and Depression: A Series of n-of-1 Trials on Effects and Feasibility. J Evid Based Complementary Altern Med. 2017;22:856–869.

16. Gaus W and Högel J. Studies on the efficacy of unconventional therapies. Problems and designs. Arzneimittelforschung. 1995;45:88–92.

17. Haas DC and Sheehe PR. Dextroamphetamine pilot crossover trials and n of 1 trials in patients with chronic tension-type and migraine headache. Headache. 2004;44:1029–1037.

18. Weng H-Y, Cohen AS, Schankin C and Goadsby PJ. Phenotypic and treatment outcome data on SUNCT and SUNA, including a randomised placebo-controlled trial. Cephalalgia: an international journal of headache. 2018;38:1554–1563.

19. Santos C and Weaver DF. Topically applied linoleic/linolenic acid for chronic migraine. J Clin Neurosci. 2018;58:200–201.

20. Margolis A and Giuliano C. Making the switch: From case studies to N-of-1 trials. Epilepsy & behavior reports. 2019;12:100336.

21. Devan H, Farmery D, Peebles L and Grainger R. Evaluation of Self-Management Support Functions in Apps for People With Persistent Pain: Systematic Review. JMIR Mhealth Uhealth. 2019;7:e13080-e13080.

22. Larsen ME, Huckvale K, Nicholas J, Torous J, Birrell L, Li E and Reda B. Using science to sell apps: Evaluation of mental health app store quality claims. NPJ Digit Med. 2019;2:18–18.

23. Larsen ME, Nicholas J and Christensen H. Quantifying App Store Dynamics: Longitudinal Tracking of Mental Health Apps. JMIR Mhealth Uhealth. 2016;4:e96-e96.

24. Grist R, Porter J and Stallard P. Mental Health Mobile Apps for Preadolescents and Adolescents: A Systematic Review. J Med Internet Res. 2017;19:e176-e176.

25. Pombo N, Garcia N, Bousson K, Spinsante S and Chorbev I. Pain Assessment--Can it be Done with a Computerised System? A Systematic Review and Meta-Analysis. Int J Environ Res Public Health. 2016;13:415–415.

26. Rosser BA and Eccleston C. Smartphone applications for pain management. J Telemed Telecare. 2011;17:308–312.

27. https://itunes.apple.com/us/app/imtracker/id1435408653?mt=8.

28. Kravitz RL, Schmid CH, Marois M, Wilsey B, Ward D, Hays RD, Duan N, Wang Y, MacDonald S, Jerant A, Servadio JL, Haddad D and Sim I. Effect of Mobile Device-Supported Single-Patient Multi-crossover Trials on Treatment of Chronic Musculoskeletal Pain: A Randomized Clinical Trial. JAMA Intern Med. 2018;178:1368–1377.

29. Vega-Barbas M, Diaz-Olivares JA, Lu K, Forsman M, Seoane F and Abtahi F. P-Ergonomics Platform: Toward Precise, Pervasive, and Personalized Ergonomics using Wearable Sensors and Edge Computing. Sensors (Basel, Switzerland). 2019;19.

30. Lamb ZW and Agrawal DP. Analysis of Mobile Edge Computing for Vehicular Networks. Sensors (Basel, Switzerland). 2019; 19.

31. Kobayashi S, Kane TB and Paton C. The Privacy and Security Implications of Open Data in Healthcare. Yearbook of medical informatics. 2018.

32. Gabriel MH, Noblin A, Rutherford A, Walden A and Cortelyou-Ward K. Data breach locations, types, and associated characteristics among US hospitals. The American journal of managed care. 2018;24:78–84.

